# Development of prediction models for antenatal care attendance in Amhara region, Ethiopia

**DOI:** 10.1101/2022.11.16.22282409

**Authors:** Bryan Wilder, Clara Pons-Duran, Frederick G. B. Goddard, Bezawit Mesfin Hunegnaw, Sebastien Haneuse, Delayehu Bekele, Grace J. Chan

## Abstract

**Background:** In low-resource settings, coverage of at least four antenatal care (ANC) visits remains low. As a first step towards enhancing ANC attendance, this study aims to develop a series of predictive models to identify women who are at high risk of failing to attend ANC in a rural setting in Ethiopia.

**Methods:** This is a cohort study conducted in the Birhan field site, Amhara region. Using data of a surveillance system and a pregnancy cohort, we developed and internally validated a series of logistic regressions with regularization (LASSO), and ensembles of decision trees.

Discrimination was estimated using the area under the receiving operator characteristic curve (AUC). Three prediction time points were considered: conception, and gestational weeks 13 and All models were internally validated using 5-fold cross validation to avoid overfitting.

**Results:** The study sample size was 2195. Mean age of participants was 26.8 years (Standard Deviation (SD) 6.1) and mean gestational age at enrolment was 25.5 weeks (SD 8.8). A total of 582 women (26.5%) failed to attend ANC during cohort follow-up. We observed AUC in the range of 0.61-0.70, with higher values for models predicting at weeks 13 and 24. All AUC values were similar with slightly higher performance for the ensembles of decision trees.

**Conclusion:** This study presents a series of prediction models for ANC attendance with modest performance. The developed models may be useful to identify women at high risk of missing their ANC visits to target interventions to improve attendance rates. This study opens the possibility to develop and validate easy-to-use tools to predict health-related behaviors in settings with scarce resources.

**SUMMARY BOX:**

- No published studies to date have developed risk prediction models for ANC attendance.
- The presented models show modest performance, but may be useful to identify pregnancies at a high risk of not initiating ANC.
- This type of models could be used by countries with strong community health programs to identify high-risk women to target specific interventions aiming to improve ANC attendance rates, increasing feasibility and cost-effectiveness of those interventions.
- Our models were internally validated using cross-validation to avoid overfitting, and despite not being tested in other populations, they are useful to inform local and regional health authorities.
- This study demonstrates that it is possible to develop predictive models for behavioral outcomes using data from surveillance systems and pregnancy cohorts in settings with scarcity of resources.

## INTRODUCTION

Antenatal care (ANC) is the provision of essential maternity services and interventions aiming to prepare the mother for the birth, to ensure the healthy evolution of the pregnancy, and to prevent, diagnose and anticipate complications.[1] Evidence shows that ANC provision has shown benefit in preventing maternal and neonatal deaths,[2-4] as well as improving birth outcomes.[5, 6] Since the 1990s, ANC attendance has been based on a model of a minimum of four visits.[1] Based on evidence suggesting that additional visits decrease perinatal mortality and increase mother’s satisfaction,[7] the WHO launched its most recent ANC guidelines in 2016, recommending eight contacts during pregnancy instead of a minimum of four. Despite the new WHO ANC model, country and regional estimates regarding ANC attendance are only available for the FANC model in most countries. In Ethiopia, the latest estimates from 2019 Ethiopia Mini Demographic and Health Survey (EMDHS) indicate that 74% of women aged 15 to 49 who had a live birth in the five years before the survey had at least one ANC visit attended by a skilled health provider. However, the percentage of those who had at least four ANC visits was lower, at 43%.[8] The proportions varied significantly across country regions, and between urban and rural areas. Urban women were more likely to attend at least four ANC visits (59%) than women living in rural areas (37%). Of note, the EMDHS estimates were all based on self-reported information.

A few studies have demonstrated that mass media and mobile health interventions may improve ANC attendance.[9-11] However, there is still a knowledge gap in evidence-based interventions to enhance ANC attendance and adherence in sub-Saharan African countries.[12] The first step towards designing and implementing strategies to encourage women to attend ANC services, is to identify those individuals who may be more prone to skip visits. To the best of our knowledge, no previous studies have developed predictive models that address this need.

It is critical to ensure universal access to quality ANC services and to identify population groups that are less likely to access those services in settings with low ANC coverage rates. Stratifying women at risk for low ANC coverage could target interventions to improve ANC attendance and to mobilize resources for populations at highest risk. In this context, we conducted a pregnancy and birth cohort with health and demographic surveillance in Amhara region, Ethiopia. Our study aims to develop a series of predictive models to identify women who are at high risk of failing to attend ANC in rural Amhara, Ethiopia, using information on factors collected at different timepoints before and during pregnancy, at community and facility level.

## METHODS

### Study design and setting

We conducted a cohort study in the Birhan field site, including 16 villages in Amhara region, Ethiopia, covering a population of 77,766, to estimate morbidity and mortality outcomes among 17,108 women of reproductive age and 8,554 children under-five with house-to-house surveillance every three months. The site is a platform for community and facility-based research and training that was established in 2018, with a focus on maternal and child health.[13] Nested in the site is an open pregnancy and birth cohort that enrolls approximately 2,000 pregnant women and their newborns per year with rigorous longitudinal follow-up over the first two years of life and household data linked with health facility information.[14] The catchment area is rural and semi-urban, covers both highland and lowland areas, and includes two different districts, Angolela Tera, and Kewet/Shewa Robit.[13]

We used data from the Birhan Health and Demographic Surveillance System (HDSS) and the nested pregnancy and birth cohort, Birhan Maternal and Child Health (MCH) cohort, to develop a series of prediction models for ANC attendance.[13, 14] The HDSS provides estimates and trends of health and demographic outcomes including morbidity among women of reproductive age and children under two years and births, deaths, marriages, and migration in the entire population.[13] The pregnancy and birth cohort generates evidence on pregnancy, birth, and child outcomes using clinical and epidemiological data at both community and health facility level.[14]

### Study population

We used data of women enrolled during pregnancy in the MCH cohort between December 2018 and March 2020. Of those, we excluded women who delivered after 9^th^ April 2020, since ANC visits were no longer recorded in the facility charts after that date due to COVID-19 pandemic restrictions in the study area. We further excluded newborns with implausible gestational ages at birth: <28 weeks due to the definition of stillbirth in Ethiopia, and ≥ 46 weeks. Gestational age was estimated using the best available method from ultrasound measurements, reported date of last menstrual period, fundal height or maternal recall of gestational age in months. Detailed information on the hierarchy of how the best available method was chosen can be found elsewhere.[15]

### Outcomes

The principal outcome of the predictive models was failing to attend at least one ANC visit during pregnancy. In the cohort, ANC visits that took place in the study catchment area during pregnancy follow-up after enrolment were captured by prospective facility chart abstraction. In a subset of participants, data collectors abstracted ANC visits from facility charts retrospectively to minimize missing data from visits that occurred prior to enrolment and were missed by data collectors. Visits with a record in the mothers’ medical chart, abstracted either retrospectively or prospectively, were used to create the outcome variable of the predictive models. Of note, some ANC visits before enrolment and those that happened outside of the study catchment area may have been missed from facility chart abstraction.

### Predictors

The initial selection of potential predictors was guided by a literature review of papers published between 1990 and 2022 reporting on factors associated with ANC attendance in Sub-Saharan Africa. We searched PubMed using the following search terms: (“prenatal” OR “antenatal” OR ANC) AND (“risk factors” OR “determinants”). Additional factors collected as part of the HDSS and the MCH cohort were also considered for inclusion. Following the removal of predictors with ambiguous definitions and low prevalence rates in the data (i.e. fewer than five cases), a range of socio-economic, demographic, medical, environmental and pregnancy-related predictors were considered for inclusion in the predictive models. The complete list of included predictors can be found in Supplemental Material 1.

### Analysis

#### Descriptive analysis

A descriptive analysis of the background characteristics of study participants was performed. Binary variables were reported using counts and percentages, whereas continuous variables were reported using median and interquartile range (IQR).

#### Prediction models

Supervised classification models were fit to predict the binary outcome of whether a woman will have at least one ANC visit during pregnancy. The positive outcome denotes that a woman had no ANC visits, as the goal was to identify women who are at risk of receiving no ANC. Two kinds of models were fit: a logistic regression model with regularization via the least absolute shrinkage and selection operator (LASSO) to avoid overfitting;[16] an ensemble of decision trees (XGBoost) which can automatically leverage nonlinear interactions between predictors.[17] All models were fit and internally validated using 5-fold cross validation.[18] Both models perform a form of variable selection during fitting, the LASSO via penalizing model coefficients towards zero and decision trees by selecting a small subset of variables to branch on.

The discrimination of all models was estimated using the area under the receiving operator characteristic curve (AUC) on the held-out data within each cross-validation fold. An AUC value of 0.5 represents a random prediction which is uncorrelated with the true outcome. Larger values indicate more accurate predictions, and a value of 1 represents predictions which are perfectly concordant with the true outcome. Receiving operator characteristic (ROC) curves were generated to assess the potential clinical utility of the models by measuring its specificity at a fixed level of sensitivity.

We compared the predictive performance that can be achieved by each model as a function of the gestational age at which a prediction is made. This measures the clinical utility of the model in a scenario where risk prediction is performed at a given point in pregnancy, using all information available up until that time point. At a given time *t*, we predicted attendance only for women who did not attend any ANC visits by *t*. This assesses the ability of the model to differentiate between women for whom it is still unknown (at *t*) whether they will receive ANC care or not. We considered three distinct time points: *t* = 0, i.e., using only information available at the time of conception, *t =* 13 weeks, and *t* = 24 weeks.

#### Missing data handling

Ensemble of decision trees automatically handle missing values for the predictors by picking a default branch in each tree along which to route observations with missing values. For logistic regression with LASSO, we performed multiple imputation via the Multivariate Imputation by Chained Equations (MICE) package for R.[19] Before performing imputation, dummy variables indicating missingness for each predictor were included as additional variables, an approach that is justified for predictive models because it reflects the complete state of knowledge available at the time of prediction.

### Ethical considerations

Ethical clearance was obtained from the Ethics Review Board (IRB) of Saint Paul’s Hospital Millennium Medical college, (Addis Ababa, Ethiopia) [PM23/274], and Harvard T.H. Chan School of Public Health (Boston, United states) [IRB19-0991]. Signed informed consent was obtained from all participants.

### Patient and public involvement

Patients and/or the general public were not involved in the design, execution or drafting of this study.

## RESULTS

The initial sample was composed of 2801 women. We first removed 73 cases (2.6%) with extreme gestational ages and 2 (0.1%) who did not have information on gestational age. Then, 475 women out of the remaining 2726 (17.4%) delivered after the onset of COVID-19 pandemic – or were expected to deliver after that date in case of censored participants – and were also excluded from the study. Of the remaining 2251, we excluded 56 women (2.5%) who were lost to follow-up before delivery and before attending any ANC visits. The final study sample size was 2195.

Mean age at conception of participants was close to 27 years (Table 1). Mean gestational age at enrolment in the cohort was 26 weeks, between second and third trimester. Almost half of the sample were women who cannot read and write (44.9%), there was an even representation of women from both districts in the study area (43.4% from Angolela Tera district), and almost a third of participants were in their first pregnancy (32.3% primiparous). The proportion of study participants who did not have any recorded ANC visits during pregnancy (prospective or retrospective) was 26.5%.

**Table 1.** Characteristics of the study sample

| Variables | Participants |  |  |
| --- | --- | --- | --- |
|  | N | mean | SD |
| Age at conception (years) | 2189 | 26.8 | 6.1 |
| Gestational age at enrolment (weeks) | 2195 | 25.9 | 8.8 |
|  | N | n | % |
| Cannot read and write | 2190 | 983 | 44.9 |
| District: Angolela Tera | 2191 | 951 | 43.4 |
| Primiparous | 2195 | 709 | 32.3 |
| No recorded ANC visits | 2195 | 582 | 26.5 |

Some predictors had high levels of missing data, especially where we relied on information collected by the health system (i.e. signs and symptoms) for the week 13 and 24 models. Not all included individuals were enrolled by weeks 13 and 24; approximately 90% and 60% of participants missed data on signs and symptoms for the respective models. Anthropometric measurements collected before pregnancy were missing for over 55% of the cohort because those were not collected in all HDSS rounds. Approximately 10% of women had missing data on nutritional habits.

Models were fit to predict the probability that each woman would attend no ANC visits, varying the time (measured in gestational age) at which the prediction was made. Table 2 shows the value of AUC with 95% confidence interval (CI) achieved for each model and prediction time. We observe AUCs in the range of 0.61-0.70, with values generally higher after the first trimester than at the start of pregnancy (i.e., using only information available at time of conception). Figure 1 shows the ROC curves for the logistic regression with LASSO and the ensemble of decision trees at each time point. ROC curves and AUC values were generally similar between the two models, with slightly higher performance for the ensemble of decision trees, though with any difference in AUC values contained within the 95% confidence interval.

**Table 2.** AUC of predictive models

| Time | Logistic regression with LASSO | Ensemble of decision trees |
| --- | --- | --- |
|  | AUC (95% CI) | AUC (95% CI) |
| Conception | 0.61 (0.58, 0.64) | 0.62 (0.59, 0.65) |
| Week 13 | 0.68 (0.66, 0.71) | 0.70 (0.67, 0.72) |
| Week 24 | 0.66 (0.64, 0.69) | 0.67 (0.64, 0.69) |

*AUC – area under the receiving operator characteristic curve; CI – confidence interval; LASSO – least absolute shrinkage and selection operator*

**Figure 1.**
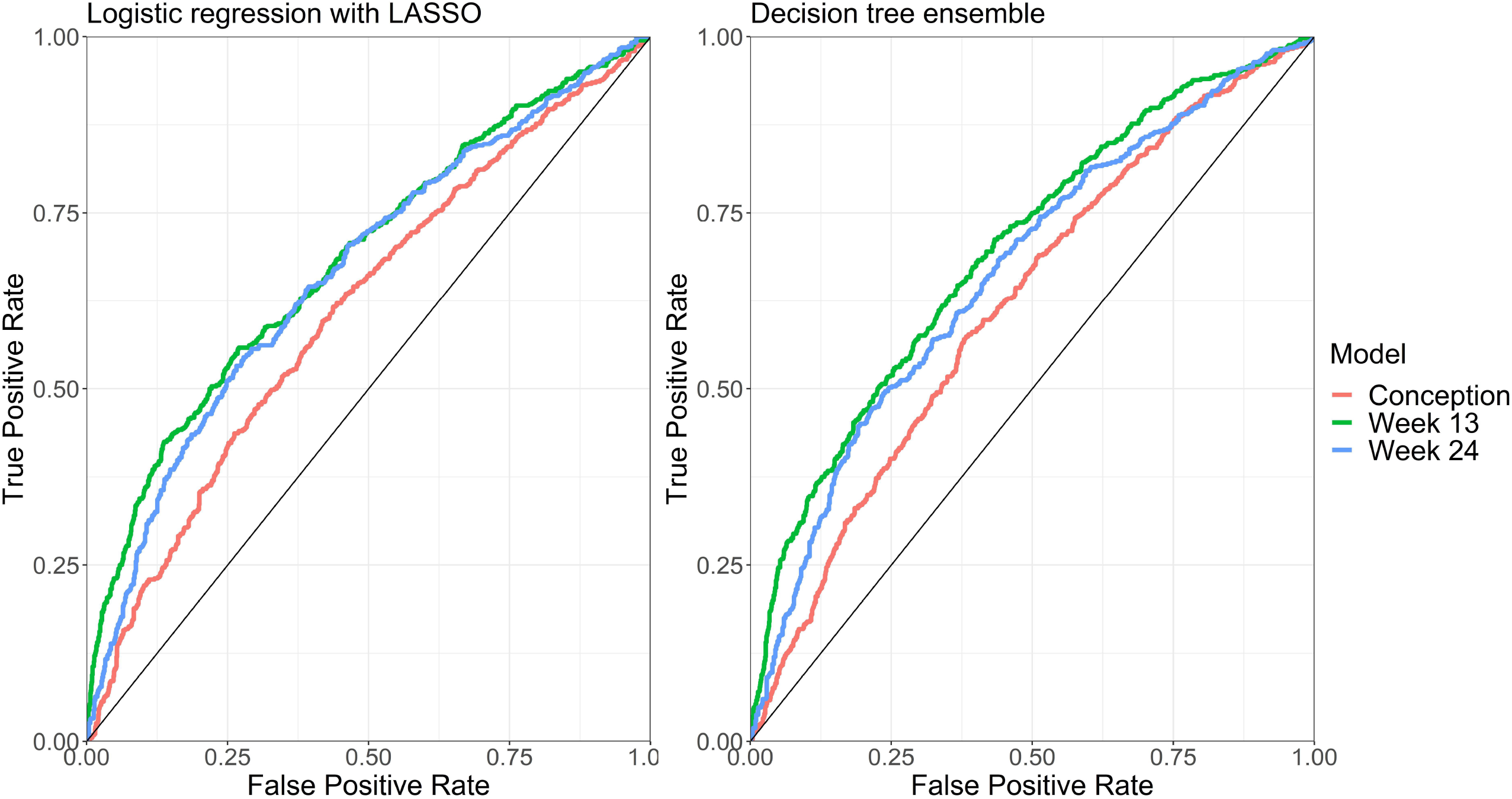
ROC curves for each predictive model and time of prediction. Footnote: LASSO - least absolute shrinkage and selection operator; ROC - receiving operator characteristic curve

Table 3 and Table 4 show the odds ratios (OR) of the logistic regression with LASSO at each time point, and the feature importance for the three ensembles, respectively, for the fifteen predictors with the strongest associations with the outcome. We observe that demographic features (e.g. age, ethnicity, religion), dietary features, income sources, use of contraception prior to pregnancy, anthropometrics, missingness of information on obstetric history, and distance to a health facility all consistently played a large role in the predictive models. At later time points (13 and 24 weeks of gestational), not receiving community health visits from study data collectors also predicted lack of facility ANC visit attendance (OR 4.05 in logistic regression with LASSO for both week 13 and 24 models).

**Table 3.** Logistic regression with LASSO: odds ratios (OR) for top predictors

| Variable | Conception<br>OR | Week 13<br>OR | Week 24<br>OR |
| --- | --- | --- | --- |
| Prior contraceptive use | 0.67 | 0.77 | 0.87 |
| Eat fortified food weekly | 0.77 | 0.7 | 0.87 |
| Education = none | 1.16 | 1.22 | 1.14 |
| Time to health facility = missing | 4.14 | 1.17 | - |
| Income source = merchant | 1.41 | 1.29 | - |
| Income source = private business | 2.09 | 1.6 | - |
| Interpregnancy interval = missing | 0.84 | 0.65 | - |
| No community visit | NA | 4.05 | 4.05 |
| Ethnic group = Oromo | 1.71 | 1.29 | 1.03 |
| Occupation = petty trade | 0.83 | 0.89 | 0.59 |
| History of baby with birth defects | 4.32 | 1.8 | - |
| Previously leaving in a different region (migrated) | 0.4 | - | - |
| Occupation = self-employed | 1.42 | 1.51 | - |
| Urinary urgency | NA | 2.39 | - |

*LASSO – least absolute shrinkage and selection operator ; NA – Not applicable; OR – odds ratio*

**Table 4.** Ensemble of decision tree: feature importance for top predictors

| Variable | Conception<br>Importance | Week 13<br>Importance | Week 24<br>Importance |
| --- | --- | --- | --- |
| Age at conception | 0.05 | 0.05 | 0.04 |
| Body mass index | 0.03 | 0.05 | 0.03 |
| Prior contraceptive use | 0.06 | 0.03 | 0.02 |
| Time living at current residence | 0.04 | 0.02 | 0.04 |
| Education = higher | 0.01 | 0.04 | 0 |
| Family size | 0.02 | 0 | 0.03 |
| Height | 0.05 | 0.03 | 0.03 |
| Distance to health facility | 0.04 | 0.03 | 0.01 |
| Time to health facility | 0.18 | 0.1 | 0.07 |
| Mid-upper arm circumference | 0 | 0.03 | 0.01 |
| Religion = Muslim | 0.04 | 0.02 | 0.01 |
| No community visit | NA | 0.25 | 0.33 |
| Age of first pregnancy | 0.01 | 0.01 | 0.03 |
| Weight | 0.08 | 0.04 | 0.01 |

NA – Not applicable

## DISCUSSION

This study demonstrates that, despite the low specificity of the models, it is possible to develop predictive models for behavioral outcomes such as ANC attendance using data from HDSS and pregnancy cohorts in a setting where there is usually lack of information on key pregnancy-related health indicators and a complete reliance on self-reports to assess health services utilization. The developed models predict with modest performance the probability that a woman from the Birhan field site does not initiate ANC during pregnancy at three different prediction times. The best performing algorithm was an ensemble of decision trees that obtained an AUC of 0.70 for the prediction at week 13 of gestation. This AUC can be interpreted as the probability that a randomly selected woman who will never attend an ANC visit is correctly classified as presenting higher risk to fail to attend ANC than a randomly selected woman who will access the services. With the available data, prediction at other timepoints is poorer.

To the best of our knowledge, this is the first study to date aiming to develop an accurate model to predict failure to access ANC services in a low-resource setting. Previous studies carried out in Ethiopia and other low-resource areas focused on assessing coverage and factors associated with ANC attendance, but none reported performance of models or internally validated those. These studies specifically looked at social, economic, and demographic factors, and found that socioeconomic status, educational level, distance from the health center or marital status among others, are associated with access to ANC services.[4, 20, 21] Obstetric and medical history related factors as well as signs and symptoms during the first weeks of gestation, are less explored in relation to attendance to ANC in Ethiopia. For instance, women with a previous history of stillbirth are less prone to attend ANC services in subsequent pregnancies.[22] In an effort to build the most accurate and well-performing models possible with available data, we used both types of predictors in our study, as well as habits and nutritional factors. Factors of different domains were found among the top predictors of failure to attend ANC in our models. In terms of habits and behaviors, women who reported use of contraceptives in HDSS rounds before getting pregnant or taking fortified foods were less likely to fail to attend ANC visits. Regarding socio-demographic factors, participants who did not know how far they live from the nearest health facility, those who get their main income from work in a private company, or who belong to Oromo ethnic group were more prone to not initiate their ANC follow-up. With regard to obstetric history, the most important predictor of failure to utilize ANC was having had a baby with birth defects. Causes of failure to attend ANC need to be further studied. Prediction studies like this aim to characterize prognosis, and a different study design and analytical approach is required to infer causality.

Despite the wide range of predictors considered, the models still show modest results that could benefit from additional data. An increase in sample size may contribute to keeping predictors that were excluded from the study due to low prevalence rates and thus, potentially improve the predictive performance of models. Additionally, we hypothesize that it may be necessary to use data on trust in the health system such as health services utilization before pregnancy, and healthcare seeking behavior like ANC attendance in previous pregnancies to enhance the predictive ability of the algorithms.

The development of prediction models for ANC attendance may have implications for future research and operational activities in countries with strong community health programs such as Ethiopia. Our data shows that not receiving pregnancy follow-up visits in the community may predict propensity to not initiate facility ANC visits. The Ethiopian Government has been implementing the Health Extension Program since 2003, where Health Extension Workers and the Health Development Army have a central role in health promotion at primary level.[23] Globally, the most common task carried out by community health workers (CHWs) is health promotion,[24] and the availability of easy-to-handle tools that allow them to identify women at high risk of not attending their visits and disregarding recommendations, may be of high impact. Although one of their responsibilities is already to encourage women to attend ANC facilities, experiences from other sub-Saharan African countries show that CHWs programs do not always translate into increased utilization rates.[25, 26] Voluntarism, high workload and burnout of certain profiles of CHWs in Ethiopia are barriers to expand their scope of work and provide additional counselling to all women at community level. [27, 28]

Prediction models with modest performance may be used to identify pregnancies at a high risk of never initiating ANC, and thus those could be targeted in specific interventions to improve ANC attendance rates. However, the lack of accuracy of our models may imply that still a considerable proportion of individuals at moderate to high risk will not be targeted in such interventions, but the strategy of intensifying efforts towards a subset of the population might be feasible and cost-effective.

### Strengths and limitations

Most ANC coverage estimates from low-resource settings, including Ethiopia, rely on self-reports. [8, 29, 30] One of the strengths of our study is the contribution of longitudinal facility recorded data to conservatively count the total number of ANC visits attended by study participants to predict ANC access. As a limitation, the counts used to generate ANC coverage may underestimate the total number of visits attended due to possible visits outside the study catchment area, or incomplete retrospective data abstraction. The potential underestimation of ANC visits may affect the results of the prediction model by conservatively predicting that women did not receive ANC care when their visits were simply unlikely to be recorded. The longitudinal design, the large sample size and the novelty in developing the first predictive models for ANC attendance, are additional strengths of this study.

Study limitations include the need to exclude pregnancies that ended after the onset of COVID-19 pandemic, thus reducing the study sample size. However, we assumed our results may be applicable to the current situation in the Birhan field site where pregnant women may behave similarly as during the pre-pandemic years. Further, the level of missing data in some of the predictors may be considered a caveat of the study. However, missing data were handled using statistical procedures (e.g. MICE for LASSO models). Lack of external validation is a weakness of many published studies on predictive model development. To our knowledge there are no existing datasets that could be used to externally validate the models. Our models were internally validated using cross-validation to reduce the probability of model overfitting due to the use of a single dataset. Future studies are needed to externally validate these models in other populations.

## Conclusion

Our study presents a series of prediction models for ANC attendance that show modest performance. Due to their low specificity, these models are most appropriate for the identification of women at particularly high risk of not utilizing ANC services and are less able to differentiate between women at moderate risk. Our study opens the possibility to start exploring the development and validation of easy-to-use tools to predict health-related behaviors in settings with scarcity of resources.

## Data Availability

Data are available upon reasonable request to the authors. Data use is governed by the Birhan Data Access Committee (DAC) and follows Birhan's data sharing policy. All researchers who wish to access Birhan data can complete a Birhan data request form and submit it for decision by the Birhan DAC. Datasets will only be provided with deidentified data to maintain confidentiality of study participants.

## AUTHOR CONTRIBUTIONS

BW, CPD, FGBG, SH and GJC conceptualized and designed the study. GJC is the study PI, she obtained funding for the study, and supervised all study activities. DB is the co-PI of the study, he helped to obtain funding for the study and led data collection team. BMH, DB and GJC participated in data collection. BW and CPD conducted the data analysis. SH oversaw the data analysis. FGBG curated the data. BW and CPD drafted the first version of the manuscript. All authors critically revised the manuscript for important intellectual content. All authors approved the final version of the manuscript.

## ACKNOWLEDGMENTS

We thank all the mothers and children who participated in the study (HDSS and MCH cohort) and the community of the Birhan field site. We also thank data collectors, supervisors, coordinators, and the HaSET team for their contributions.

## SUPPLEMENTAL MATERIALS

Supplemental Material 1. Table S1. Predictors included in the models

Supplemental Material 2. TRIPOD Checklist

