## Supplemental File 1 for "Development of prediction models for antenatal care attendance in Amhara region, Ethiopia"

**Table S1. Predictors included in the models**

| **Predictors** | **Models by time point of prediction** | | |
| --- | --- | --- | --- |
|  | **Conception** | **Week 13** | **Week 24** |
| **Socioeconomic and demographic** | | | |
| Maternal age | X | X | X |
| Maternal education | X | X | X |
| Maternal literacy | X | X | X |
| Marital status | X | X | X |
| Occupation | X | X | X |
| Family’s main source of income | X | X | X |
| Family wealth index | X | X | X |
| Family size (number of individuals in the household) | X | X | X |
| Ethnicity | X | X | X |
| Religion | X | X | X |
| Woreda | X | X | X |
| Time and distance to the nearest health facility | X | X | X |
| Previous residence | X | X | X |
| Time she has been living in study area | X | X | X |
| **Anthropometric measurements** | | | |
| Preconception weight | X | X | X |
| Height | X | X | X |
| Preconception body mass index | X | X | X |
| Preconception mid-upper arm circumference | X | X | X |
| **Environmental and behavioral** | | | |
| Individual frequency of tv and radio use | X | X | X |
| Individual ownership of mobile phone | X | X | X |
| Alcohol consumption | X | X | X |
| Khat consumption (stimulant drug) | X | X | X |
| Nutritional habits (frequency of intake of several products) | X | X | X |
| Availability of food to feed children in last 30 days | X | X | X |
| Availability of food to feed herself in last 30 days | X | X | X |
| Use of contraceptives before pregnancy | X | X | X |
| **Medical and obstetric history** | | | |
| Past medical history of diabetes | X | X | X |
| Past medical history of sexually transmitted diseases | X | X | X |
| History of stillbirth | X | X | X |
| History of miscarriage | X | X | X |
| History of preterm birth | X | X | X |
| History of multiple gestation | X | X | X |
| History of C-section | X | X | X |
| History of low birth weight newborn | X | X | X |
| History of baby with birth defects | X | X | X |
| Gravidity | X | X | X |
| Parity | X | X | X |
| Interpregnancy interval | X | X | X |
| Location of last delivery | X | X | X |
| Age at first pregnancy | X | X | X |
| Last pregnancy was planned | X | X | X |
| Experience of past infant deaths | X | X | X |
| Last child born is still alive | X | X | X |
| **Signs and symptoms during pregnancy** | | | |
| Vomiting and severe nausea |  | X | X |
| Headache |  | X | X |
| Blurry vision |  | X | X |
| Right upper quadrant abdominal pain |  | X | X |
| Urinary pain |  | X | X |
| Increased urinary frequency |  | X | X |
| Urinary urgency |  | X | X |
| Decreased fetal movement |  | X | X |
| Vaginal bleeding |  | X | X |
| Iron and folic acid supplementation |  | X | X |
| **Operations-related** | | | |
| Community visits by study field workers during pregnancy |  | X | X |
